# High Demand, Low Possession: Dilemmas and Strategies for Research Capability Cultivation in Clinical Medicine Postgraduates

**DOI:** 10.64898/2026.06.12.26355561

**Authors:** Boyang Wang, Lulu Yang, Zhaohui Gong

## Abstract

Most previous studies have examined medical postgraduate research training from a single dimension, lacking a full-chain analysis that integrates capability demand, actual possession, obstacles, and output. Consequently, the measurement of capability gaps and the analysis of underlying training model deficiencies remain insufficient. To address this gap, we administered a self-designed multidimensional questionnaire to 86 clinical medicine postgraduates at a medical school, covering research cognition, interest, capability demand and possession, participation pathways, difficulties, and outputs. The aim was to systematically characterize the current situation, identify problems, and propose optimization strategies. Over 90% of participants expressed interest in research, yet only 1.16% self-rated as very knowledgeable. The largest demand-possess gap was for writing and publication (86.05% vs. 16.28%), followed by independent research capability (75.58% vs. 11.63%). A total of 59.30% cited lack of foundational knowledge, making experiments very difficult, as the greatest challenge, and 66.28% had no research achievements. The primary source of research topics was supervisor assignment (54.65%), with only 4.65% choosing topics independently. No statistically significant differences were found across grades or training types (*P* > 0.05). These findings reveal a structural high demand, low possession gap in medical postgraduate research training, with early research experience deficit and a passive research model as key constraining factors. Accordingly, an integrated bachelor–postgraduate progressive research competency training system is proposed.

## 1 Introduction

Research innovation capability is at the core of postgraduate education. Medicine, combining the attributes of natural sciences and clinical practice, undergoes rapid knowledge updates, with diagnostic and therapeutic breakthroughs relying on research support. The “Guidance of the General Office of the State Council on Accelerating the Innovative Development of Medical Education” emphasizes “cultivating high-level medical talents with a spirit of research innovation,” elevating research innovation capability to a national strategic level[1]. However, the current cultivation of research capabilities among clinical medicine postgraduates faces multiple challenges: under enrollment expansion, students come from diverse backgrounds with varying research foundations; professional degree programs emphasize clinical work, compressing research time, while academic degree postgraduates experience dual pressures from in-depth thesis projects and graduation requirements. Medical postgraduates generally exhibit high research interest but weak capabilities, high participation but low output[2–4]. Surveys show that only 21.1% of postgraduates self-assess as having independent research capability[5]; insufficient training in research thinking and paper writing is the primary bottleneck[6].

Most previous studies are limited to a single dimension, lacking a full-chain examination from “capability demand – actual possession – obstacles – output.” There is insufficient measurement of capability gaps and analysis of the causes of training models. In view of this, this study takes currently enrolled postgraduates at a medical school as subjects, employing a multidimensional questionnaire covering research cognition, interest, capability demand and current status, participation pathways, difficulties, and outputs. The goal is to systematically present the current situation, diagnose problems, and propose optimization recommendations.

## 2 Methods

### 2.1 Study design and participants

This study used convenience sampling to select currently enrolled postgraduates at Ningbo University. Ethical approval for this study was obtained from the institutional review board at Ningbo University Health Science Center under the reference number NBU-20231005. A total of 90 questionnaires were distributed, and 86 valid questionnaires were returned, yielding an effective response rate of 95.56%. The basic characteristics of the respondents are shown in **Table 1**.

**Table 1.** Basic characteristics of participants (n=86)

| Item | Value | Number of students | Percentage (%) |
| --- | --- | --- | --- |
| Gender | Male | 41 | 47.67 |
|  | Female | 45 | 52.33 |
| Year | PGY 1 | 39 | 45.35 |
|  | PGY 2 | 33 | 38.37 |
|  | PGY 3 | 14 | 16.28 |
| Training type | Academic | 48 | 55.81 |
|  | Professional | 38 | 44.19 |

PGY, postgraduate year.

### 2.2 Questionnaire design and analysis

Based on a review of existing literature [2–6] and pre-interviews with five senior postgraduates, we self-developed the “Questionnaire on Research Innovation Capabilities of Medical Postgraduates.” The questionnaire contains 20 items covering the following dimensions: (1) basic information (gender, year, training type); (2) research cognition and interest (pre-admission understanding, current interest level); (3) demand for and self-rated possession of research capabilities (10 capability indicators: literature reading, question posing, problem analysis, problem solving, writing & publication, independent research, communication, coordination & balance, stress resistance, self-discipline); (4) research participation experience (undergraduate participation, timing of first participation, source of research topic); (5) research difficulties and achievements (main difficulties, existing achievements; types of achievements were multiple-choice); (6) evaluation of environment and satisfaction (supervisor competence, hardware environment, academic atmosphere, overall self-assessment); (7) open-ended suggestions (fill-in-the-blank). The questionnaire was reviewed by three medical education experts (two professors and one associate professor, each with over 10 years of postgraduate supervision experience); the average content validity index (S-CVI/Ave) was 0.89. After a pilot survey of 20 students, the internal consistency was tested, and Cronbach’s α coefficient was 0.82, indicating acceptable internal consistency reliability.

### 2.3 Data processing and statistical analysis

Data entry and statistical analysis were performed using SPSS 26.0 software. Descriptive statistical methods were mainly used, with categorical data presented as frequencies and percentages. Differences between capability demand and possession were compared, and the “demand rate – possession rate” difference was calculated to quantify capability gaps. To explore capability differences among groups, chi-square (χ^2^) tests were used to compare differences in the possession rates of core capabilities across grades (first-year/second-year/third-year) and training types (academic degree/professional degree).

## 3 Results

### 3.1 Weak research cognition: high interest, low goals

The survey showed that medical postgraduates had a very weak foundation of research cognition before enrollment. Only 1.16% indicated “very knowledgeable” about research, 55.81% “moderately knowledgeable,” and 43.02% “uninformed.” This indicates that over 98% of students had not established a systematic understanding of research. However, the interest level was relatively optimistic. In total, over 90% of students held a positive attitude toward research. In terms of goal setting, only 47.67% hoped to “produce high-level achievements,” while 41.86% only aimed to “meet graduation requirements,” with over 40% anchoring to the minimum standard. Thus, although students have a strong interest in research, they generally lack foundational knowledge and high-level aspirations.

### 3.2 Insufficient higher-order output capabilities and a large supply-demand gap

This study focused on ten core research capabilities, asking postgraduates to select items they “believe they need” and items they “believe they currently possess.” The results are shown in **Table 2**.

**Table 2.**
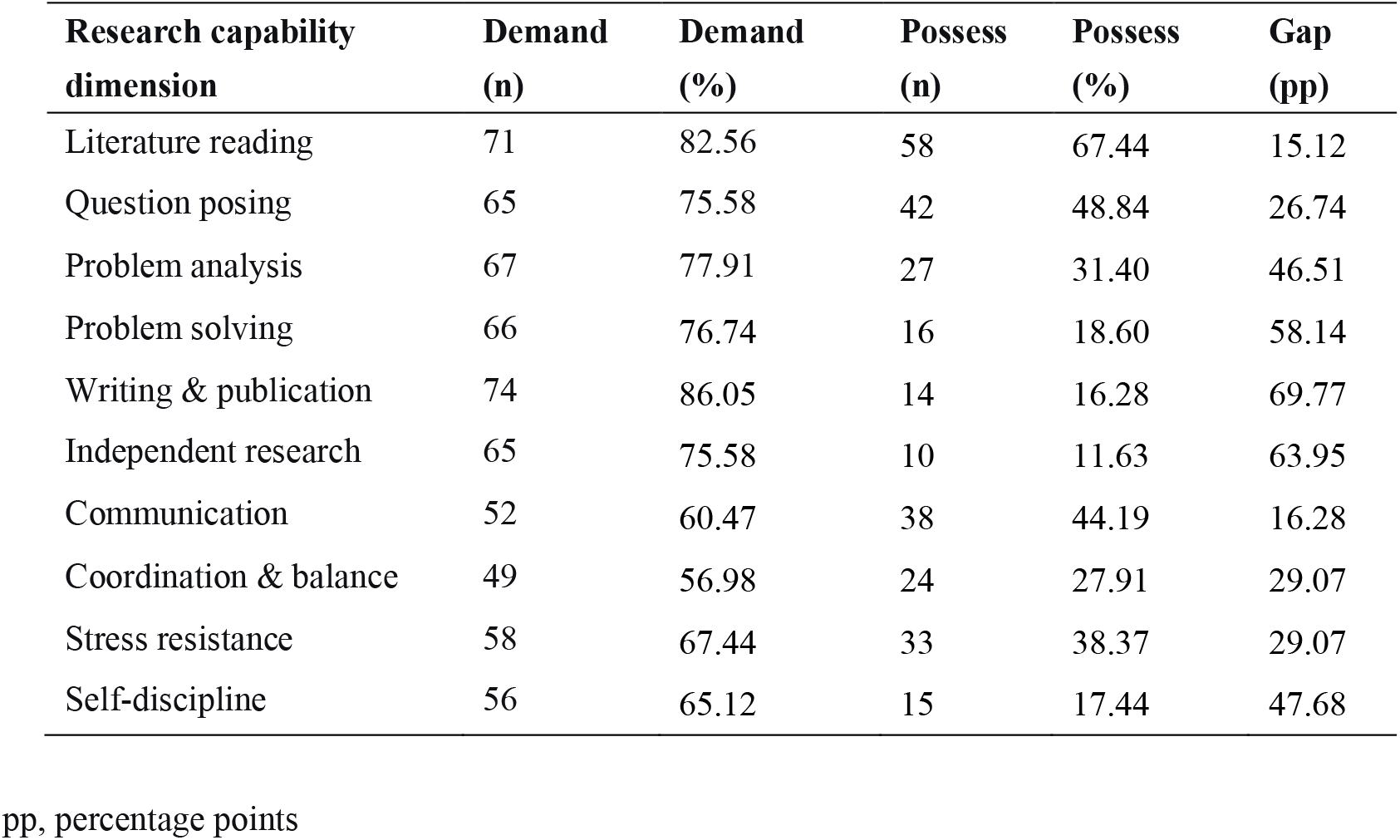
Comparison of demand for and self-rated possession of research capabilities among medical postgraduates (n=86)

From the demand side, the top five capabilities considered most necessary by postgraduates were: writing and publication (86.05%), literature reading (82.56%), problem analysis (77.91%), problem solving (76.74%), and question posing and independent research (both 75.58%), highlighting the high importance placed on research output and full-process capabilities. In contrast, the capabilities with relatively high possession rates were only literature reading (67.44%), question posing (48.84%), and communication (44.19%). Most critically, the supply-demand gap was largest for paper writing and publication (69.77 percentage points (pp)), followed by independent research (63.95 pp), problem-solving (58.14 pp), strong self-discipline (47.68 pp), and problem analysis (46.51 pp). This gap profile clearly reveals that while students generally master the basic “input-oriented” skill of literature reading, they are severely deficient in “output-oriented” higher-order abilities such as writing, independent research, and problem solving.

### 3.3 Low undergraduate research participation and high dependence on supervisors for topic selection

The results showed that undergraduate research participation was low. Only 31.40% of students had participated in research activities during college, while 68.60% had never participated. Correspondingly, the timing of first research participation was highly concentrated in “PGY 1 of postgraduate studies” (59.30%), with only 25.58% having participated in early or late undergraduate years, and 11.63% indicating they had “never participated” to date. This suggests that for most students, the postgraduate stage is the starting point of research. Regarding topic sources, supervisor-assigned topics dominated (54.65%), topics determined through student-supervisor consultation accounted for 36.05%, and topics chosen based on personal interest accounted for only 4.65%, with 4.65% selecting “other.” This result reflects that current medical postgraduates’ research topic selection is highly dependent on supervisor arrangements, with very limited student autonomy.

### 3.4 Weak foundation leading to research difficulties, and few achievements attributed to insufficient personal effort

Regarding the greatest difficulty currently encountered in conducting research, the top choice was “lack of foundation, making experiments very difficult,” accounting for 59.30%. This finding corroborates the earlier observations of weak research cognition and lack of early experience. The second and third greatest difficulties were “inability to balance life and work” (15.12%) and “high project difficulty, insufficient supervisor guidance” (8.14%). In contrast, “low supervisor competence, arbitrary direction” (4.65%) and “inadequate laboratory hardware conditions” (6.98%) accounted for relatively low proportions.

Overall, the current state of research achievements is not optimistic (**Table 3, 4**). 66.28% (57 students) had no research achievements of any form (including those in progress).

**Table 3.** Research achievements already obtained *(n=86)

| Achievement type | As first author (n, %) | As co-author (n, %) |
| --- | --- | --- |
| Published papers | 8 (9.30%) | 4 (4.65%) |
| Granted patents | 3 (3.49%) | 1 (1.16%) |
| Scientific wards | 4 (4.65%) | 2 (2.33%) |

\*Some students had multiple achievements, so the total number of individuals does not equal the sum of items.

**Table 4.** Research achievements in progress (n=86)

| Achievement type | As first author (n, %) | As co-author (n, %) |
| --- | --- | --- |
| Manuscript in preparation/submission/under review | 7 (8.14%) | 3 (3.49%) |
| Patents or awards pending | 2 (2.33%) | 1 (1.16%) |

When asked to identify the primary factor influencing research achievements, 53.49% of students selected “personal effort,” 24.42% attributed it to “supervisor competence,” 11.63% to “laboratory hardware conditions,” and 10.47% to “other.” Students generally exhibited an internal attribution tendency, closely linking achievement output to personal effort.

### 3.5 Positive perceptions of supervision and environment, but low self-rated research satisfaction

The evaluation of supervisor competence was generally positive. 67.44% of students considered their supervisor’s competence “high, fully meeting guidance needs,” 26.74% “moderate, basically meeting guidance requirements,” and only 3.49% “poor.” Regarding guidance frequency, 58.14% indicated “frequent,” 32.56% “moderate, able to discuss with the supervisor when needed,” and a total of 9.30% considered guidance “infrequent” or “almost none.” For laboratory hardware environment, 48.84% rated it as “moderate, basically meeting research requirements,” 36.05% as “good, fully meeting requirements,” and 15.11% as “poor” or “uncertain.” The academic atmosphere was more favorable: 59.30% rated it as “good, many opportunities for exchange, harmonious relationships,” 25.58% as “moderate, less exchange,” and none selected “poor.”

In overall self-assessment, only 3.49% expressed being “very satisfied” with their current research status, 10.47% “satisfied,” 56.98% “moderate,” while the combined proportion of “dissatisfied” and “very dissatisfied” was 22.09%, and 6.98% were “uncertain.” The ratio of satisfaction to dissatisfaction was approximately 1:2, indicating a generally low self-rating of research status among students.

### 3.6 Diverse research gains: cognitive and capability growth as primary

The greatest gains from engaging in research were diverse (multiple-choice question). The top three were: “improved understanding of medical scientific issues” (73.26%), “recognized my own shortcomings” (68.60%), and “enhanced ability to conduct independent research” (60.47%). These were followed by “improved teamwork ability” (50.00%) and “gained friendships” (32.56%). Only 5.81% explicitly stated “no gains.” This indicates that even when research output is limited, most students still obtain positive experiences in cognitive improvement, self-reflection, and capability growth from the process.

### 3.7 Observed differences in research capabilities across grades and training types

To further explore factors influencing research capabilities, this study compared the possession rates of core capabilities among different grades and training types. Regarding grade differences, third-year students had a higher possession rate of “paper writing and publication ability” (28.57%, 4/14) than first-year (10.26%, 4/39) and second-year students (15.15%, 5/33), but the difference was not statistically significant (χ^2^=2.84, *P*=0.242). Similarly, the possession rates of “independent research capability” across the three grades were 7.69% (first-year), 12.12% (second-year), and 21.43% (third-year), with no statistically significant difference (χ^2^=2.11, *P*=0.348). Regarding training type differences, academic degree postgraduates had a higher possession rate of “independent research capability” (16.67%, 8/48) than professional degree postgraduates (5.26%, 2/38), but the difference was not significant (χ^2^=2.68, *P*=0.102). The possession rates of “paper writing and publication ability” between academic degree (18.75%, 9/48) and professional degree (13.16%, 5/38) postgraduates also showed no significant difference (χ^2^=0.49, *P*=0.484).

## 4 Discussion

### 4.1 Structural fracture in research capability cultivation: the input-output chasm

The most striking finding of this study is the large gap between the demand for and actual possession of research capabilities among medical postgraduates, particularly the most pronounced disconnects in “paper writing and publication ability” (demand 86.05% vs. possession 16.28%, gap 69.77 percentage points) and “independent research capability” (demand 75.58% vs. possession 11.63%, gap 63.95 percentage points). This is consistent with previous research: Li et al. found that paper writing ability is a common shortcoming among medical postgraduates[7]; Bakhshi et al. noted that insufficient research training is a key barrier to research output[8]. The underlying cause is a tendency in the current curriculum to “emphasize input over output.” Most medical schools offer courses in literature retrieval and statistics in the first postgraduate year, enhancing literature reading and comprehension ability (possession rate 67.44% in this study), but systematic training in academic writing and independent project design is lacking. Students have to “learn by doing” without organized instructional design. As Castillo-Martínez stated, academic writing ability is crucial for the sustained innovation of higher education systems[9]. Furthermore, the capability gap also manifests as a broken “analysis–solution” chain: students’ self-rated possession rate for “question-posing ability” was 48.84%, which dropped to 31.40% for “problem-analysis ability,” and further to 18.60% for “problem-solving ability,” showing a decreasing trend. It should be noted that these three abilities were each self-rated by students on different items of the same questionnaire, so common method bias is possible. The observed decreasing trend can only preliminarily suggest a capability dilemma of “being able to identify problems but struggling to systematically analyze them, being able to analyze problems but struggling to solve them independently,” which awaits validation with more refined measurement tools.

### 4.2 Passive research model constrains innovation capability formation

In this study, 54.65% of research topics came from “supervisor assignment,” while only 4.65% were independently chosen by students. Although supervisor-assigned topics have their rationale – supervisors using their academic experience to provide direction for new students – when this becomes the dominant model, its negative impact on innovation capability cannot be ignored. The essence of research innovation lies in discovering new problems, proposing new hypotheses, and designing new solutions. If students are placed in a passive position from the very starting point of topic selection, subsequent research often degenerates into technical execution of the “supervisor’s will” rather than independent exploration. When students perceive the topic as “the supervisor’s” rather than “mine,” their depth of engagement and persistence may suffer. Research has shown that autonomy support can stimulate higher learning motivation and self-fulfillment[10]. The data from this study indirectly support this argument: although 67.44% of students acknowledged their supervisor’s competence and only 4.65% felt that supervisor guidance was insufficient, research output remained low (66.28% had no achievements). This indicates that even high-quality and frequent supervisor guidance does not automatically translate into students’ independent research capability. Capability generation requires agentic practice – experiencing a complete research cycle in a project where one “takes the lead and takes responsibility.”

### 4.3 Lasting effect of early research experience deficit

This study found that nearly 70% of students (68.60%) had not participated in research activities during their undergraduate years, and 59.30% began their first research experience only in the first year of postgraduate studies. There is a clear causal chain between this “zero-starting-point” reality and the fact that 59.30% of students identified “lack of foundation, making experiments very difficult” as the greatest difficulty. The value of undergraduate research participation lies in “demystifying” research – understanding basic processes, building expectations for setbacks, and forming reasonable expectations of supervisor–student relationships. Missing this stage, students often face a triple discomfort in cognition, skills, and psychology upon entering postgraduate studies. Our research group’s previous work found that early research training is significantly correlated with innovation capability[11]; Shanahan et al. demonstrated that undergraduate research experience is a significant positive predictor of postgraduate research self-efficacy and persistence[12]; Fu et al. also found that undergraduate research training has a facilitating effect on development at the postgraduate stage[13]. More concerning, the absence of early research experience may lead to a “catch-up trap”[14]: students need to simultaneously compensate for foundational skills, establish research thinking, and produce output during the postgraduate period, and the stacking of multiple tasks exacerbates anxiety. When students struggle to get started due to a weak foundation, supervisors may tend to provide more specific technical guidance rather than thinking training, thereby compressing students’ space for independent thought – forming a vicious cycle of “weak foundation → passive learning → insufficient thinking training → slow capability improvement.”

### 4.4 Dual implications of attributing outcomes to personal effort

In this study, 53.49% of students attributed the primary factor influencing research achievements to “personal effort.” This strong internal attribution tendency has a dual meaning. On the one hand, it reflects students’ sense of responsibility and agency, which aligns with the autonomous qualities required for research work. On the other hand, when 66.28% of students have no achievements and 59.30% feel a “poor foundation,” it would be simplistic to conclude that this is simply a result of students “not working hard enough.” A more reasonable interpretation is that the training system lacks supportive mechanisms to effectively convert “personal effort” into “research capability” and “research output.” In other words, students are not lacking effort; rather, they lack the scaffolding and stepping stones to make that effort productive. Therefore, the key to reforming medical postgraduate education is not simply to encourage students to “work harder,” but to provide systematic, targeted support for capability development so that effort can be effectively deployed.

## 5 Proposed strategies and recommendations

Based on the above findings and discussion, this paper proposes the following optimization pathways for cultivating research innovation capability among medical postgraduates.

### 5.1 Building an integrated bachelor–postgraduate progressive research training system

The fundamental solution to the “zero-starting-point” predicament of medical postgraduates is to shift the starting point of research capability cultivation forward to the undergraduate stage. Recommendations: (1) Add compulsory courses on research methods in the senior years of clinical medicine undergraduate programs, covering literature search and evaluation, fundamentals of research design, and principles of common experimental techniques, so that students possess basic research cognition and skills before entering postgraduate studies; (2) Establish an undergraduate research mentorship system, encouraging undergraduates to join faculty research groups for observational learning and auxiliary work, thereby demystifying research; (3) At the postgraduate stage, establish “Progressive Research Capability Workshops” that follow the full-process sequence of “literature review → project design → experiment implementation → data analysis → paper writing → submission and revision,” systematically and organizedly enhancing students’ comprehensive research capabilities.

### 5.2 Establishing student-led open funds to expand topic autonomy

To break the single model of passively receiving topics, it is recommended to establish a “Postgraduate Independent Exploration Open Fund.” The features of this fund are: small project scale, short duration (6–12 months), flexible funding, with the primary goal of training independent research capability. With supervisor consent and guidance, students independently pose scientific questions, design plans, and write applications, which are then funded after evaluation by a panel of college experts. The value of such “open projects” is not in producing high-level outputs, but in enabling students to experience the full cycle of research management – “posing questions → designing plans → applying for resources → implementing research → summarizing and reporting” – thereby cultivating their sense of agency as independent researchers and project management skills. This emphasizes that learners construct knowledge and capabilities through active inquiry.

### 5.3 Establishing a writing–research–reflection training mechanism

Targeting the largest capability gap revealed by this study, it is recommended to systematize and advance paper writing training. Specific measures include: (1) In the second semester of the first postgraduate year, offer a mandatory “Medical Academic Writing” workshop employing a dual-feedback mechanism of “peer review + supervisor feedback,” improving writing ability through iterative revision; (2) Require students to complete at least one “Research Reflection Report” per semester in group meetings, systematically reviewing and analyzing their gains, confusions, and methodological reflections at that stage, thereby training problem-analysis and problem-solving abilities; (3) Encourage supervisors, in their guidance, to shift from “providing answers” to “asking good questions,” using Socratic questioning to stimulate students’ independent thinking rather than directly offering solutions.

### 5.4 Refining research evaluation and incentive orientation, and strengthening process-based incentives

The current research achievement evaluation centered on “paper publication” can easily cause frustration for postgraduates who are in the capability-building phase. It is recommended to add process-based evaluation indicators to graduation requirements, such as “quality of research progress reports,” “presentation performance in group meetings,” and “normativity of lab records,” while reducing the weight of single output indicators. At the same time, establish process-oriented awards such as “Research Progress Award,” “Best Research Design Award,” and “Outstanding Research Reflection Report Award,” so that students receive timely recognition for capability improvements at different stages. According to self-efficacy theory, accumulation of successful experiences is the most important source of enhancing self-efficacy and sustaining behavioral persistence[15]. When students receive positive feedback and recognition at multiple points in the research process, their research confidence and sustained motivation will be effectively strengthened.

## 6 Conclusions

Through a questionnaire survey, this study has revealed the current landscape and core problems of research innovation capability among postgraduates at a medical university. The main conclusions are as follows. First, the cultivation of research innovation capability for medical postgraduates faces a structural dilemma of “high demand, high willingness, low possession, low output.” Students have strong research interest and strongly perceive the need for various capabilities, but their actual possession is severely insufficient, especially the largest supply-demand gaps in paper writing and publication ability and independent research capability. Second, the capability cultivation gap has its model-based roots. Lack of early research experience leads to a “zero-starting-point” entry into postgraduate studies; the passive topic-acceptance model weakens agency and experience of the complete research process; and the “input-over-output” tendency in the curriculum results in the absence of systematic training in higher-order capabilities. Third, solving the dilemma requires systematic reform. Shifting the starting point of research capability cultivation forward to the undergraduate stage, expanding students’ topic autonomy, establishing a closed-loop training mechanism of writing–independent research–reflection, and optimizing evaluation and incentive orientation are four integrated key measures.

This study has some limitations. The sample came from only one medical university with a relatively small size (n=86), limiting generalizability. The survey used self-ratings, so the possession levels of capabilities may be subject to self-report bias. The cross-sectional design cannot reveal the dynamic process of capability development. Future research could expand the sample scope, combine supervisor evaluations and objective output data for multi-dimensional measurement, and employ longitudinal designs to explore the developmental trajectory and key influencing factors of research capability. Nevertheless, this study has systematically outlined the current dilemmas in cultivating research innovation capability among medical postgraduates. The proposed “capability gap analysis framework” and four training pathways can provide preliminary reference and practical direction for postgraduate education reform in medical universities.

## Data availability statement

The original contributions presented in the study are included in the article, further inquiries can be directed to the corresponding author/s.

## Ethics statement

The studies involving humans were approved by Ethical Review Board of Ningbo University Health Science Center (NBU-20231005). The studies were conducted in accordance with the local legislation and institutional requirements. Written informed consent for participation in this study was provided by the participants.

## Author contributions

BW: Writing–original draft, Methodology, Investigation, Formal analysis, Data curation. LY: Writing–original draft, Methodology, Investigation, Data curation. ZG: Writing–review and editing, Formal analysis, Data curation, Conceptualization, Funding acquisition, Supervision.

## Acknowledgements

The authors appreciate medical undergraduates and postgraduates at Ningbo University Health Science Center for their participating in anonymous surveys and feedback sessions.

